# The potential of attractive insecticide-treated nets (ITNs) in reducing malaria transmission: a modeling study

**DOI:** 10.1101/2025.04.02.25325102

**Authors:** Nicolas Moiroux, Cédric Pennetier

**Affiliations:** MIVEGEC, IRD, CNRS, Univ. Montpellier - Montpellier, France; Pôle de Zoologie Médicale, Institut Pasteur Dakar - Dakar, Sénégal

**Keywords:** ITN, LLIN, vector control, behavior, plasmodium falciparum, resistance

## Abstract

**Introduction:** Recent studies suggest that insecticide-treated nets (ITNs) may actively attract malaria vectors, increasing their likelihood of coming into contact with the insecticide while potentially reducing personal protection. The impact of such attractive ITN on malaria transmission at the community level remains unclear. In this study, we developed a model to compare the effectiveness of attractive, inert and deterrent ITNs under varying levels of human usage and different degrees of physiological or behavioral resistance to insecticides in malaria vectors.

**Methods:** We developed a model to simulate the host-seeking, feeding and mortality (HSFM) of mosquito vectors facing ITNs. This model allows mosquitoes to choose between two rooms based on the presence and remote influence (attractive, inert or deterrent) of ITNs. The HSFM model was then integrated into a malaria transmission model to compare the *Plasmodium* transmission potential of mosquitoes exposed to these different type of ITNs under various scenarios of ITN coverage, physiological resistance, quantitative behavioral resistance, and qualitative behavioral resistance. Results: The model predicts that attractive ITNs consistently reduce malaria transmission potential of vectors more effectively than inert or deterrent ITNs, even in the presence of resistant vector phenotypes. For instance, at intermediate use rate (50%), strongly attractive ITNs are expected to reduce transmission by up to 67% compared to deterrent ITNs. In resistant vector populations, attractive ITNs remained more effective overall, though the reduction in transmission were less pronounced.

**Conclusion:** Our findings suggest that both inert and attractive ITNs could enhance malaria control efforts, outperforming current deterrent ITNs, even in resistant vector populations. Shifting from deterrent to inert or attractive ITNs could significantly improve vector control strategies, warranting further research and product development to fully explore and optimize this approach.

## Introduction

Malaria is a parasitic disease caused by *Plasmodium* protozoa transmitted between humans by *Anopheles* mosquitoes. In 2022 alone, an estimated 249 million cases of malaria and 608 000 deaths were reported worldwide [1]. Thanks to Insecticide-Treated Nets (ITNs), the malaria targets of the 2015 Millennium Development Goals were achieved. Between 2000 and 2015, an estimated 663 million cases were averted, with 68% of this reduction attributed to ITNs [2]. During this period, ITNs were the most widespread intervention, with usage rates increasing globally from less than 2% to 54% [1].

However, despite this progress, malaria remains a serious public health concern. In fact, while the number of deaths remains stable, global malaria cases have actually been rising since 2014, with an increase of 19 million cases in 2022 compared to 2014 [1]. Furthermore, sub-Saharan Africa continues to bear more than 90% of the disease burden, with most of the reported cases occurring in this region [1].

Several factors may be contributing to this resurgence, including insufficient ITN coverage and usage, and the spread of insecticide resistance mechanisms among malaria vector populations [3]. Resistance to pyrethroids - the only insecticide class used in ITNs until recently – has become widespread in sub-Saharan African [4]. The prevalence of both physiological and behavioral resistant phenotypes has increased among malaria vector populations, allowing mosquitoes to survive or avoid contact with treated ITNs, thereby enabling continued transmission to persist. Physiological resistance mechanisms, which enhance mosquito survival after ITN contact, are the most extensively studied. Knockdown resistance (*kdr)* mutations and the overexpression of detoxification enzymes have been well characterized, both of which reduce mosquito susceptibility to pyrethroids [5]. In contrast, behavioral resistance mechanisms are less studied but can be classified as qualitative or quantitative [6]. Qualitative behavioral resistance mechanisms allow mosquitoes to avoid insecticide contact through spatial, temporal, or trophic change. Examples include change in biting time or increased exophagy following implementations of ITN or indoor residual spraying (IRS) [7,8]. Quantitative behavioral resistance, on the other hand, occurs after insecticide contact and involve behavioral responses that reduce insecticide absorption. A well-documented example is the irritant effect of certain pyrethroids [9], which induces escape behaviors and decreases contact duration.

ITNs are mosquito bed nets made of polyester, polyethylene, or polypropylene fibers that are either coated with pyrethroid insecticides or have the insecticide incorporated into the fibers during manufacture. A person sleeping under an ITN acts as a lure for mosquitoes, which are intended to be killed upon contact with the insecticide-treated fibers. Thus, ITNs operate as a “pull-to-kill” vector control strategy. However, paradoxically, most WHO-recommended ITNs exhibit repellent properties, reducing mosquito attraction to human dwellings and hosts. This property, known as deterrence, is measured in experimental huts and defined as the reduction in mosquito entry rates into a hut with a human under an ITN compared to a control hut with a human under an untreated net [10].

While deterrence increases the personal protection provided by ITNs by reducing the number of bites, it also lowers the likelihood of mosquitoes contacting the insecticide, thereby decreasing mosquito mortality rates. The impact of deterrence on community-wide malaria transmission has been evaluated using mathematical models [11], which theoretically predict that although deterrent ITNs offer better personal protection, they are less effective than inert ITNs in reducing malaria transmission.

Conversely, field studies have shown unexpectedly high levels of mosquito attraction to human hosts protected by certain WHO-recommended ITNs, with greater numbers of mosquitoes collected in huts with ITNs than in control huts with untreated nets [12]. These ITNs, which enhance mosquito attraction to hosts, could potentially increase bite exposure, reducing personal protection under sub-optimal usage conditions or when nets are physically deteriorated. Vector control strategies are designed and evaluated primarily for their community-level effects and “attractive” ITNs could substantially influence both vector mortality and disease transmission dynamics. Understanding the impact of the remote (attractive, inert or deterrent) effect of ITNs at the community level is therefore essential for guiding product development and public health policy. Extrapolating findings of Killeen *et al*. [11], whose model did not account for the possibility that ITNs might increase mosquito attraction, we hypothesize that attractive ITNs could contribute to reducing malaria transmission, offering opportunities for novel ITN designs that may help address recent malaria resurgences.

In order to explore the theoretical potential of attractive ITNs to reduce malaria transmission in communities, we developed a modelling framework that combines a host-seeking, feeding and mortality (HSFM) model for vectors with a malaria transmission model. The HSFM model allows *Anopheles* mosquitoes to choose between two human hosts depending on the presence of ITNs and their remote effects (attractive, inert or deterrent). By integrating the HSFM model into a transmission model (adapted from Glunt *et al*. [13]), we compared the *Plasmodium* transmission potential by mosquitoes exposed to attractive, inert, or deterrent ITNs. Our analysis explored a wide range of realistic conditions that represent major sources of residual malaria transmission, including different ITN usage rates and varying levels of physiological resistance (ITN-induced mortality), quantitative behavioral resistance (indoor escaping behaviors), and qualitative behavioral resistance (spatial-temporal avoidance of ITNs). To our knowledge, this is the first study to explicitly model implications of attractive ITNs in terms of malaria transmission.

## Methods

### Host-seeking, feeding and mortality (HSFM) model

The HSFM model (Figure 1) is a static and deterministic model of the adult *Anopheles* feeding cycle with three physiological states (host-seeking HS, blood-fed BF, and resting R). From this model, we calculated transition probabilities among physiological states, which were then used in the transmission potential model.

**Figure 1.**
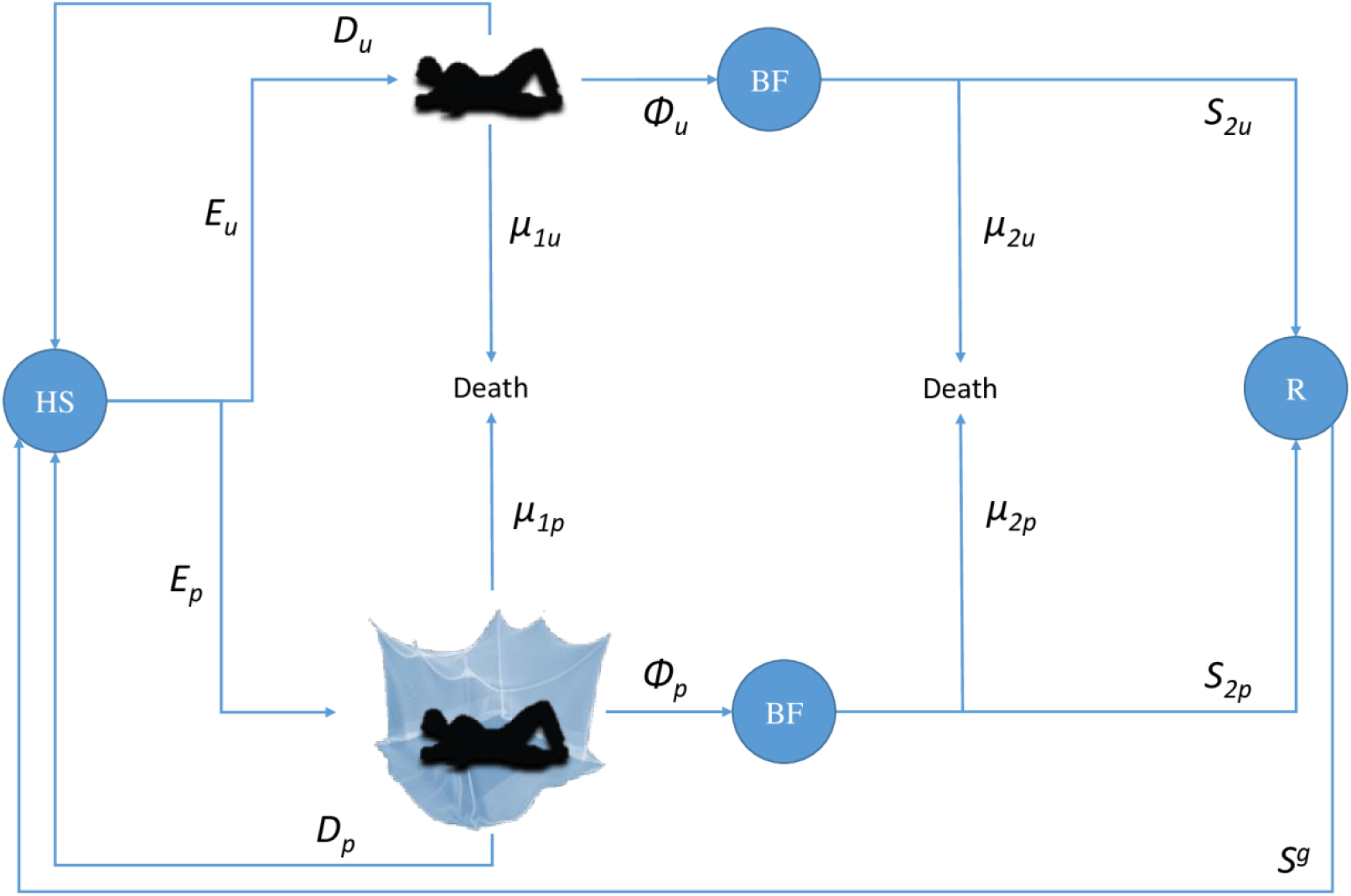
Anopheles Host-Seeking, Feeding, and Mortality (HSFM) model used to feed a malaria transmission model. Blue circles represent physiological states of Anopheles mosquitoes (HS: Host-seeking; BF: Blood-fed; R: Resting). An Anopheles vector looking for a blood-meal (in state Host-Seeking: HS) will either enter the room of an ITN-protected human (with probability E_p_) or the room of an unprotected one (with probability E_u_), depending on the remote effect of the ITN (attractive, inert or repulsive) and the human ITN usage rate in the community. After entering a hut, a vector will either be diverted (impaired and postponed to host-seeking the next night, with probability D) or attempt to feed, with two possible outcomes: feeding (with probability φ_1_) or death (with probability µ_1_). Then, blood-fed (BF) vectors will either die due to post-bite mortality (µ_2_) or survive (S_2_), entering the Resting state (R). A vector that enters state R will complete its gonotrophic cycle (g) with probability S^g^ and enter state HS to begin a new cycle.

The model considers a community of humans of known size *N*_*h*_ in which a known proportion *U*_*h*_ is using an ITN with a known ‘true’ protective efficacy of Π (i.e. the proportion of normal exposure to mosquito bites upon humans lacking ITNs, which occurs indoors at times when nets would normally be in use [11]). The average number *N*_*p*_ of people protected by an ITN is therefore:

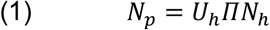

And the average number *N*_*u*_ of unprotected people is:

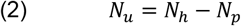

An individual *Anopheles* vector in the host-seeking (HS) state (Figure 1) in this human community will either enter the room of an ITN-protected human or that of an unprotected one, depending on the remote effect of the ITN (attractive, inert or repulsive) and the relative availability of each host type (ITN-protected or unprotected). We expressed the remote effect of ITNs as the mosquito preference (*p*_*ITN*_) for an ITN-protected host over an unprotected one. This parameter could be measured in a dual choice olfactometer (i.e. *p*_*ITN*_ would be the proportion of *Anopheles* mosquitoes that choose the ITN-protected human over the total number of *Anopheles* having made a choice) or in experimental hut trials (i.e. *p*_*ITN*_ would be the proportion of *Anopheles* mosquitoes that entered the hut equipped with an ITN over the sum of *Anopheles* mosquitoes that entered the hut with an ITN or the control hut with an untreated net). Thus:

- if *p*_*ITN*_ > 0.5, the ITN is attractive, i.e. it increases host attraction,
- if *p*_*ITN*_ = 0.5, the ITN is inert (neither attractive nor repulsive), i.e. it does not interfere with host attraction,
- if *p*_*ITN*_ < 0.5, the ITN is repulsive, i.e. it reduces host attraction,

We assumed that a host-seeking *Anopheles* mosquito is always faced with two potential hosts, each of whom may be protected or unprotected, and that it must choose between them. It cannot avoid the choice, continue searching, or switch to a non-human host. Under this assumption, the mosquito encounters one of the following cases, each with its associated probability:

- (a) two ITN-protected hosts, with probability *C*_*pp*_ or
- (b) one ITN-protected and one unprotected host, with probability *C*_*pu*_ or
- (c) two unprotected hosts, with probability *C*_*uu*_.

Probabilities *C*_*pp*_, *C*_*pu*_ and *C*_*uu*_ depend on the numbers *N*_*u*_ and *N*_*p*_ of, respectively, unprotected and ITN-protected hosts in the community. Assuming that users and non-users of ITNs are randomly distributed in space, probabilities *C*_*pp*_, *C*_*pu*_ and *C*_*uu*_ follow a hypergeometric distribution with parameters *N* = *N*_*h*_, *K* = *N*_*p*_, and *n* = 2. The hypergeometric distribution describes the probability of getting *k* successes in *n* draws without replacement from a population of size *N* containing exactly *K* successes. We calculated the probabilities that *k* = 2, *k* = 1, and *k* = 0 corresponding to cases (a), (b), and (c), respectively, using the following formula:

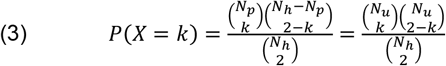

Therefore, probabilities *C*_p*p*_, *C*_p*u*_ and *C*_*uu*_ are calculated as follow:

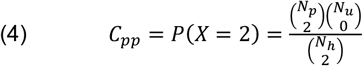

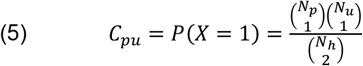

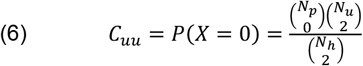

With *C*_*pp*_+ *C*_*pu*_ + *C*_*uu*_ = 1.

From this, we can compute the probabilities *E*_*u*_ and *E*_*p*_ that a host-seeking vector will enter a house with an unprotected or an ITN-protected human, respectively, as follows:

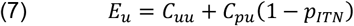

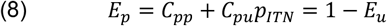

After entering a room with an unprotected host, a vector will either be diverted (impaired and postponed to host-seeking until the next night) with probability *D*_*u*_, or it will attempt to bite (with probability 1-*D*_*u*_). If it attempts to bite, the vector will either be killed (pre-bite mortality) with probability *μ*_1*u*_, or it will take a blood meal with probability *ϕ*_1*u*_, thereby entering state blood-fed (BF). Knowing *D*_*u*_ and *μ*_1*u*_, we deduce *ϕ*_1*u*_ and calculate *ϕ*_*u*_, the overall probability of successful feeding when entering a room with an unprotected host:

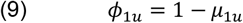

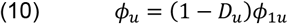

Then, vectors that blood-fed on an unprotected host will either die with probability *μ*_2*u*_ (post-bite mortality) or survive and enter the resting state (R) with probability *S*_2*u*_. Knowing *μ*_2*u*_, we have:

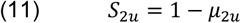

The parameters *μ*_1*u*_, *D*_*u*_, *ϕ*_*u*_, *ϕ*_1*u*_, *S*_2*u*_ and *μ*_2*u*_ have their equivalents *μ*_1*p*_, *D*_*p*_, *ϕ*_*p*_, *ϕ*_1*p*_, *S*_2*p*_ and *μ*_2*p*_, respectively, for mosquitoes entering a room with an ITN-protected host. The following expressions (12)-(14) correspond to expressions (9)-(11), but for a room with an ITN-protected host:

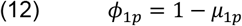

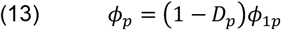

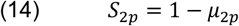

From this HSFM model described above, we then derived the variables needed to feed the “Vector average lifetime infectious bites” model of Glunt *et al*. [13], as described below.

First we calculate the probability *P*_*d*_ that a host-seeking (HS) vector will fail to feed, become diverted, and postpone its feeding attempt until the next night (conditional on surviving). *P*_*d*_ is the sum of the probabilities that a mosquito will be diverted in rooms with ITN-protected and unprotected hosts:

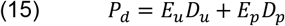

Diverted mosquitoes will survive until the next night with probability *S*_*d*_ which is equal to the baseline daily survival rate *S* of *Anopheles*:

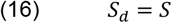

Next we calculate the probability *P*_*f,u*_ that an HS vector will successfully bite an unprotected human during the same night and enter the blood-fed state (BF). *P*_*f,u*_ is the product of the probability

*E*_*u*_ of entering a room with an unprotected host and the probability *ϕ*_*u*_ of feeding in that room:

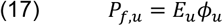

Similarly, the probability *P*_*f,p*_ that an HS vector will successfully bite an ITN-protected human during the same night is obtained by multiplying the probability *E*_*p*_ of entering a room with an ITN-protected host by the probability *ϕ*_*p*_ of feeding in that room:

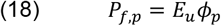

The sum of *P*_*f,u*_ and *P*_*f,p*_ gives the probability *P*_*f*_ that a host-seeking (HS) vector will successfully bite during the same night:

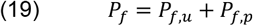

The proportion *F*_*u*_ of mosquitoes that fed on unprotected hosts is:

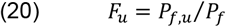

According to the HSFM model, a proportion of blood-fed (BF) mosquitoes die shortly after feeding, while the remainder survive to enter the resting (R) state (Figure 1), with probabilities *S*_2*u*_ or *S*_2*p*_ depending on whether they fed on an unprotected or ITN-protected host. Until the end of the gonotrophic cycle (of duration *g*), mosquitoes survive at a constant daily rate *S*. Therefore the probability *S*_*f*_ of surviving from a successful feeding to the next gonotrophic cycle in the host-seeking state (HS) is:

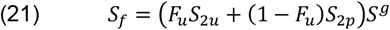

### Transmission potential (TP) model

Variables calculated above from the HSFM model are used in the model described by Glunt *et al*. [13] to estimate the transmission potential, defined as the average number of infectious bites a vector is expected to give during its lifetime. The following formulae differ slightly in notation from those used by Glunt *et al*..

First we need to determine *P*_*fA*_, the probability that an HS vector survives to take a feed. Theoretically, a vector may experience zero to an infinite number of diversions before feeding. Therefore, *P*_*fA*_ is given by a geometric series with first term *P*_*f*_ (the probability of successfull biting during the same night) and ratio *P*_*d*_*S*_*d*_ (the probability of being diverted and surviving to HS state the following night):

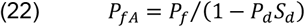

Next, we need the average number *B*_*A*_ of subsequent bites that an HS vector survives to give. *B*_*A*_ is given by a geometric series with first term *P*_*fA*_ (the probability that an HS vector survives to take a feed) and ratio *S*_*f*_*P*_*fA*_ (the probability of taking a feed and surviving to the following gonotrophic cycle in the HS state):

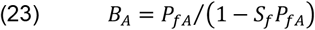

The probability *c* that a vector will become infected by *Plasmodium falciparum* while taking a blood-meal is equal to the product of the infectiousness *k* (i.e., the probability of acquiring infection while biting an infected human) and the prevalence *I*_*h*_ of *Plasmodium falciparum* infection in the human population:

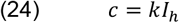

Therefore, the probability *P*_*I*_ that a vector will acquire *Plasmodium falciparum* during its lifetime is a geometric series with first term *P*_*fA*_*c* (the probability that an HS vector survives to take a feed and acquire *Plasmodium falciparum*) and ratio *S*_*f*_*P*_*fA*_(1 −*c*) (the probability of taking a non-infectious feed and surviving to the next gonotrophic cycle in the HS state):

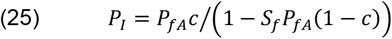

Considering a newly infected vector, it will experience a combination of diversions (*i*) and gonotrophic cycles (of duration *g*) during the sporogonic period (of duration *n*) until becoming an infectious vector. As demonsrated by Glunt *et al*. [13], the probability *S*_*I*_ that a newly infected vector will survive to the HS state as an infectious individual is given by the following equation:

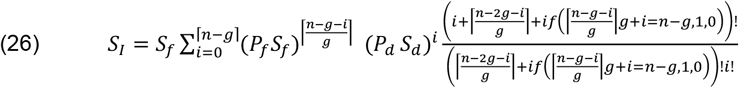

We finally computed the transmission potential *TP* (i.e., the average number of infectious bites during a vector’s lifetime, denoted *VLAIB* in [13]) by multiplying the probability *P*_*I*_ that a vector acquires *Plasmodium falciparum* during its lifetime, the probability *S*_*I*_ that a newly infected vector survives to the HS state as an infectious individual, and the average number *B*_*A*_ of subsequent bites a HS vector is expected to survive to deliver:

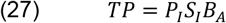

We calculated the relative transmission potential (*RTP*) between two scenarios as the ratio of the transmission potential *TP*_*test*_ computed for the tested scenario to the transmission potential *TP*_*ref*_ computed for the reference scenario:

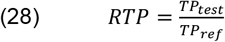

Finally, the percentage change in malaria transmission potential (*RedTP*) between tested and reference scenarios was computed based on *RTP*:

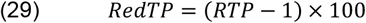

### Parameterization

The models have 15 input parameters, whose default values were defined according to the literature (Table 1). The default pre-bite and post-bite mortalities in both types of rooms (with or without ITNs), as well as diversion probability in a room with an ITN, were set to the mean values calculated from experimental hut trials in [12] (Supplementary figure 2). Because the diversion probability in a hut without an ITN (*D*_*u*_) is expected to be overestimated in recent experimental hut trials (as these used untreated holed nets as reference), we extracted values from older trials carried out during the 1980s [14,15], where unprotected peoples (i.e., with no nets at all) were used as the control. From these data, the diversion probability *D* was calculated as the proportion of unfed and alive *Anopheles* mosquitoes out of the total number of mosquitoes entering the hut. Pre-bite mortality *µ*_*1*_ was calculated as the proportion of unfed and dead *Anopheles* mosquitoes out of those that were not diverted. Post-bite mortality probability *µ*_*2*_ was calculated as the proportion of dead, blood-fed *Anopheles* mosquitoes out of all blood-fed *Anopheles* (alive or dead). Regarding other entomological parameters, we set the baseline daily survival rate *S* (during resting and after diversion) to 0.9, and the duration of the gonotrophic cycle (*g*) to 3 days, based on field data for *Anopheles gambiae* [16–18]. The default value for the duration of sporogony (*n*) was set to 11 days, corresponding to the value given by Detinova’s formula [19] at a temperature of 26°C for *Plasmodium falciparum*. The infectiousness (*k*) of mosquitoes when biting an infected human was set to 0.1 according to modelling work by Churcher *et al*. [20]. The default ITN use rate in the human population (*U*_*h*_) was set to 50%, corresponding to the current ITN use rate in sub-Saharan Africa [1]. The default proportion of human exposure to bites that occurs while ITNs are in use (Π) was set to 0.75, an intermediate value among those recorded in field studies across Africa [21–24]. We set the default value for the human community size (*N*_*h*_) to 1000 inhabitants and the prevalence of infection (*I*_*h*_) in this population to 50%, a common infection level in sub-Saharan Africa [25].

**Table 1:** Baseline parameters used in Host-Seeking, Feeding and Mortality (HSFM) and transmission models.

| Symbol | Definition | Default value | Range [min, max] or values used in simulations | Range [min, max] and distribution used in sensitivity analysis | Refs.* |
| --- | --- | --- | --- | --- | --- |
| $S$ | Daily survival rate (*An. gambiae s.l.*) | 0.9 | - | [0.61, 0.98]; uniform | [16] |
| $g$ | Average duration of gonotrophic cycle (days) | 3 | - | [2, 5]; discrete uniform | [17,18] |
| $p_{ITN}$ | Preference of vector for ITN-protected human (against unprotected human) as it might be recorded in a dual choice olfactometer or in experimental hut trials. If $p_{ITN} > 0.5$, ITN is attractive; if $p_{ITN} = 0.5$, ITN is inert (neither attractive nor repulsive); if $p_{ITN} < 0.5$, ITN is repulsive. | 0.5, 0.36 | 0.5, 0.6, 0.7 | - | [12,26] |
| $\mu_{1u}$ | Pre-bite mortality probability when attempting to bite an unprotected human | 0.03 | - | [0, 0.05]; uniform | [12] |
| $\mu_{1p}$ | Pre-bite mortality probability when attempting to bite an ITN-protected human | 0.72 | [0, 1] | [0.22, 1]; uniform | [12] |
| $D_u$ | Diversion probability when entering a room of an unprotected human | 0.12 | - | [0, 0.3]; uniform | [14,15] |
| $D_p$ | Diversion probability when entering a room of an ITN-protected human | 0.31 | [0, 1] | [0, 0.51]; uniform | [12] |
| $\mu_{2u}$ | Post-bite mortality probability after a successful feeding on an unprotected human | 0.005 | - | [0, 0.028]; uniform | [12] |
| $\mu_{2p}$ | Post-bite mortality probability after a successful feeding on an ITN-protected human | 0.29 | - | [0, 1]; uniform | [12] |
| $N_h$ | Size of the human community (N inhabitants) | 1000 | - | [300, 5000]; uniform | - |
| $I_h$ | Probability of *Plasmodium falciparum* infection in the human population | 0.5 | - | [0.1, 0.9]; uniform | [25] |
| $k$ | Infectiousness: probability that a vector become infected (exposed) while taking a blood meal on an infectious host | 0.1 | - | [0.02, 0.2]; uniform | [20] |
| $n$ | Duration of extrinsic incubation period of *Plasmodium falciparum* (days) | 11 | - | [8, 15]; discrete uniform | [19,27,28] |
| $U_h$ | ITN usage probability in the human population | 0.5 | [0, 1] | [0.1, 0.9]; uniform | [25] |
| $\Pi$ | Proportion of normal exposure to mosquito bites upon humans lacking ITNs, which occurs indoors at times when nets would normally be in use (= ITN protective efficacy) | 0.75 | [0, 1] | [0.45, 0.9]; uniform | [21–24] |

\*for default values and/or ranges used in sensitivity analysis.

### Simulations

Relative transmission potential between attractive and inert ITNs, or between the latter and deterrent ITNs, was simulated while varying (i) ITN use rate (*U*_*h*_) in the human population, (ii) mosquito pre-bite mortality (*µ*_*1p*_) in rooms equipped with ITN (representing different levels of physiological resistance), (iii) mosquito diversion (*D*_*p*_) when entering a room equipped with an ITN (representing varying levels of quantitative behavioral resistance), and (iv) the proportion (Π) of human exposure to bites that occurs while ITNs are in use (representing varying levels of mosquito spatiotemporal avoidance of ITNs, i.e., qualitative behavioral resistance).

Attractive ITNs induced vector preferences of *p*_*ITN*_ = 0.6 or *p*_*ITN*_ = 0.7, meaning that 60% or 70% of malaria vectors, respectively, selected a human under an ITN when given a choice against an unprotected human. These levels of attraction have been observed multiple times in field studies (Supplementary Figure 1) [12,26]. The inert ITN induced a vector preference of *p*_*ITN*_ = 0.5, indicating no preference (i.e., vectors were equally likely to select an ITN-protected or unprotected human). In contrast, the deterrent ITN induced a vector preference of *p*_*ITN*_ = 0.36, meaning that only 36% of malaria vectors choose a human under an ITN when presented against an unprotected human. This value represent the mean deterrence effect observed among significantly deterrent ITNs reported in the literature (Supplementary Figure 1) [12].

Relative transmission potential between deterrent, inert or attractive ITNs and no nets (i.e. 0% use) was also simulated in the same scenarios as above.

### Uncertainty and sensitivity analysis

We conducted an uncertainty and sensitivity analysis to evaluate the robustness of the model results and the impact of input parameters beyond the specific scenarios tested in prior simulations. Based on the literature, we defined plausible ranges of values for the parameters (Table 1 and Supplementary Figure 2). We then employed Latin Hypercube Sampling [29] to generate 500 simulations comparing moderately attractive ITNs to deterrent ITNs. We assumed that parameter values were sampled from uniform probability distributions, either discrete or continuous. We calculated partial rank correlation coefficients (PRCC) between outcomes and parameters, as well as partial slope coefficients (PSC; i.e., linear regression coefficients) with their 95% bootstrapped confidence intervals.

#### Software

Models were coded in R [29]. The uncertainty and sensitivity analysis was performed using the ‘LHS’ function from the ‘pse’ package version 0.4.7 [30].

## Results

We simulated the impact of providing human communities with attractive ITNs on malaria transmission potential, expressed as the average number of infectious bites a vector is expected to give during its lifetime, and compared the results to communities receiving either inert or deterrent ITNs.

### Effect of ITN usage rates

According to our model, attractive ITNs are expected to reduce malaria transmission more effectively than inert or deterrent ITNs, regardless of usage levels. With all other parameters set to default values based on literature and field data (Table 1 and Supplementary Figure 2), the greatest transmission reduction was observed at use rates of 92–94%. At these levels, a highly attractive ITN (*p*_*ITN*_ = 0.7) is expected to reduce *Plasmodium* transmission potential, expressed as the average number of infectious bites a vector delivers during its lifetime, by 56% compared to an inert ITN (Figure 2A) and by 74% compared to a deterrent ITN (Figure 2B). Similarly, at 92-94% use rate, moderately attractive ITNs (*p*_*ITN*_ = 0.6) are predicted to reduce malaria transmission by 33% and 60% compared to inert and deterrent ITNs, respectively (Figure 2A and 2B). At 50% ITN use, inert and attractive ITNs are expected to reduce transmission by 33-63% compared to deterrent ITNs (Figure 2B). Compared to no nets (0% use), attractive LLINs used by 50% of the population are predicted to reduce transmission by 93%, which is 12 percentage points (p.p.) more than deterrent ITNs (81% reduction; supplementary Figure 3A).

**Figure 2.**
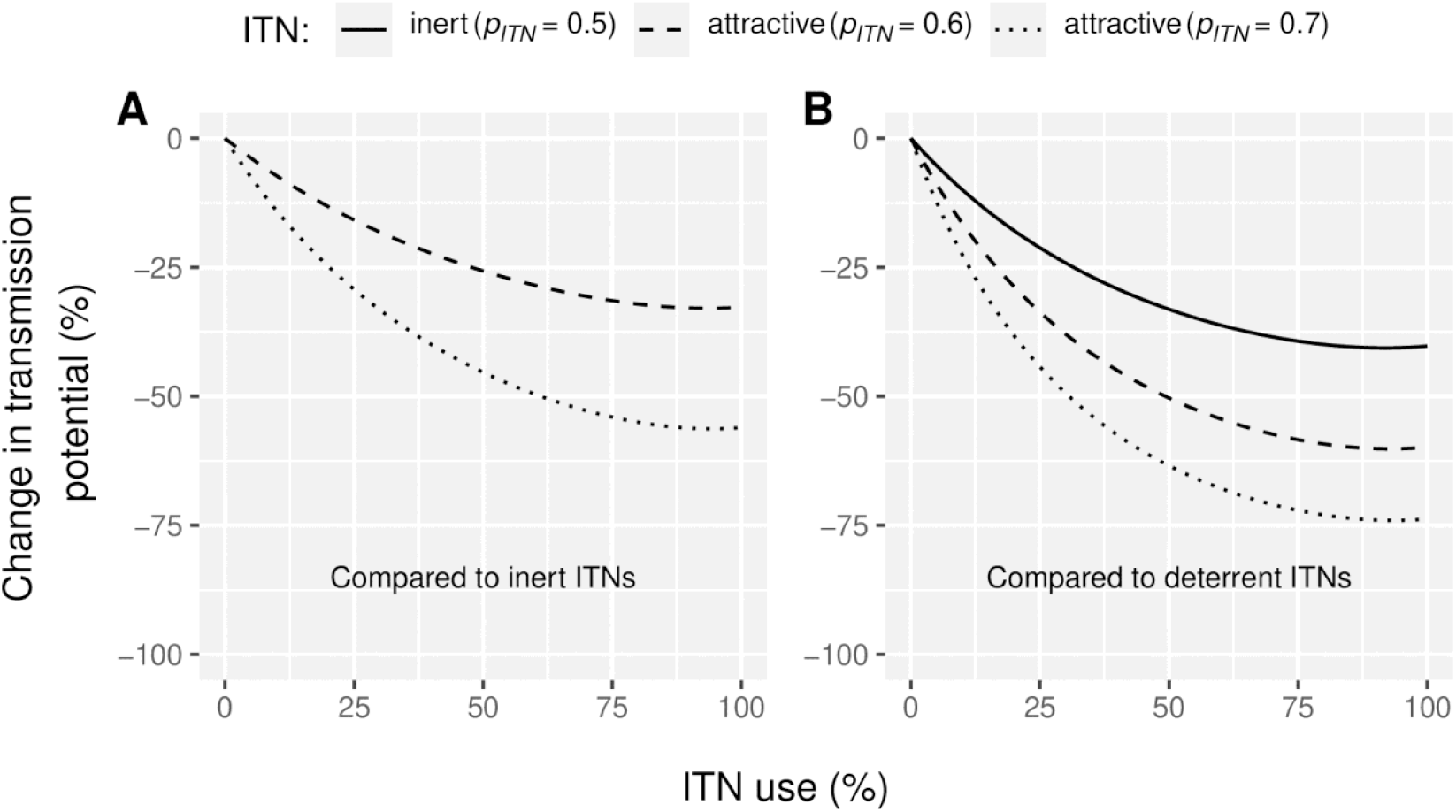
Reduction in *Plasmodium* transmission potential (*RedTP*) induced (A) by attractive Insecticide-Treated Nets (ITNs) compared to inert ITNs and (B) by attractive and inert ITNs compared to deterrent ITN for varied levels ITN usage rates in the human population (*U*_*h*_). *Vector preference p*_*ITN*_ *of 0*.*6 indicates that 60% of malaria vectors choose an ITN-protected human if presented against an unprotected human. The reference inert ITN (panel A) induces a vector preference of 0*.*5 (vectors are equally likely to select an ITN-protected or unprotected human) and the reference deterrent ITN (panel B) induces a vector preference of 0*.*36 (only 36% of the vectors choose an ITN-protected human against an unprotected human)*.

### Effect of physiological resistance

Physiological resistance was defined as the vector’s ability to survive a feeding attempt in a dwelling equipped with an ITN. In our model, the probability of survival during a feeding attempt corresponds to the likelihood of successfully taking a blood meal, conditioned on not being diverted (Figure 1). Our simulations indicate that, regardless of the level of physiological resistance in the vector population, attractive ITNs consistently outperform inert and deterrent ITNs in reducing malaria transmission (Figure 3), although the reduction in transmission declines as resistance levels increase. In the absence of physiological resistance (i.e., 0% survival), and with all other parameters held constant (including ITN use rate at 50%; Table 1), moderately attractive ITNs (*p*_*ITN*_ = 0.6) and highly attractive ITNs (*p*_*ITN*_ = 0.7) are predicted to reduce malaria transmission by 32% and 55%, respectively, relative to an inert (*p*_*ITN*_ = 0.5) ITN (Figure 3A). When compared to a deterrent ITN (*p*_*ITN*_ = 0.36) under the same conditions (no resistance), an inert ITN is expected to reduce malaria transmission by 41%, while moderately attractive (*p*_*ITN*_ = 0.6) and highly attractive ITNs (*p*_*ITN*_ = 0.7) are expected to reduce transmission by 60% and 73%, respectively (Figure 3B). Under high resistance conditions (82% survival i.e. the highest level observed in field data [12]; supplementary Figure 2B), both moderately and highly attractive ITNs still provide better community protection but yield moderate reductions in transmission potential: of 13% and 25%, respectively, relative to inert ITNs (Figure 3A) and of 29% and 38%, respectively, relative to deterrent ITNs (Figure 3B). Compared to no nets (0% use), attractive LLINs (50% use) are predicted to achieve a 74% reduction in transmission potential under high resistance conditions (82% survival), which is 17 percentage points (p.p.) more than deterrent ITNs (supplementary Figure 3B).

**Figure 3.**
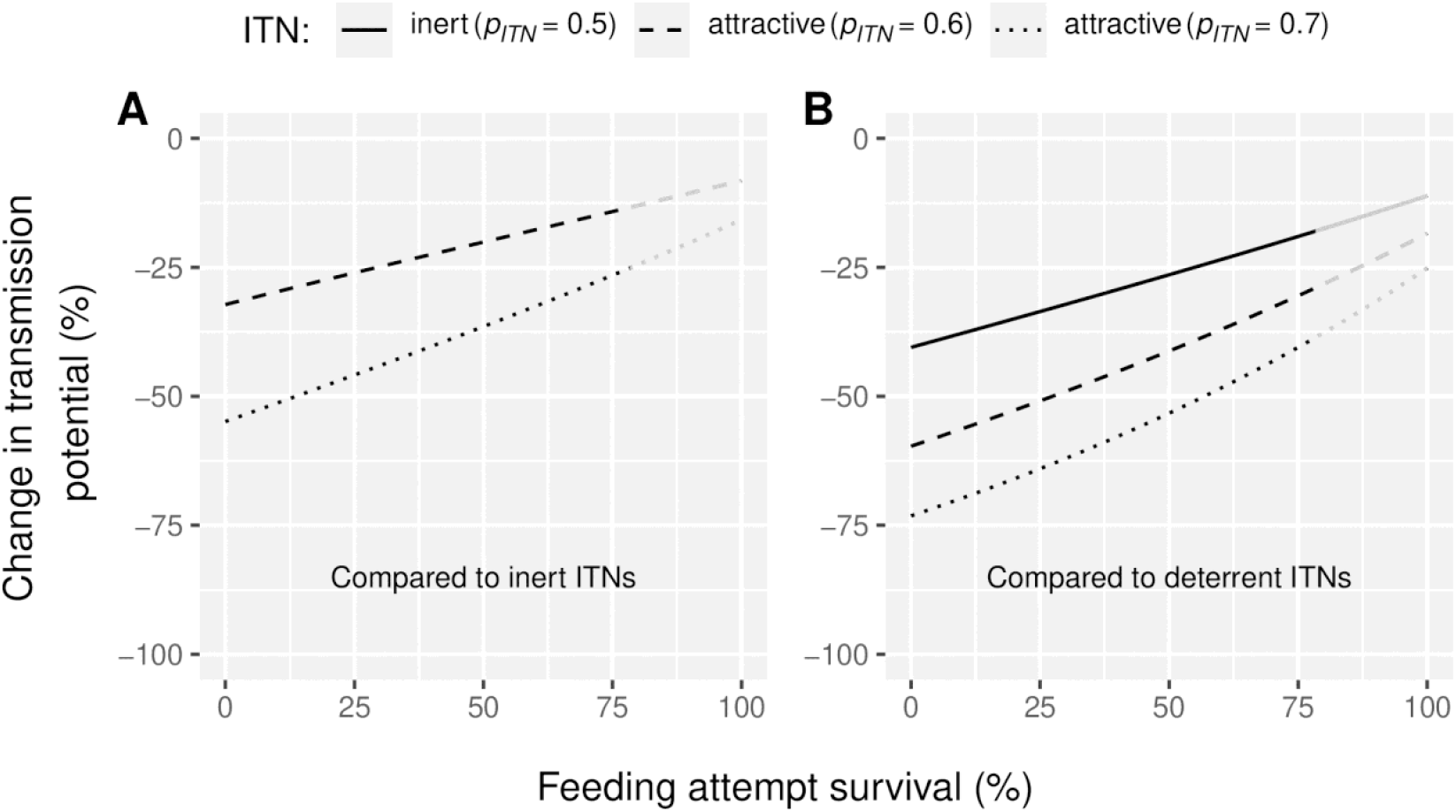
Reduction in *Plasmodium* transmission potential (*RedTP*) induced (A) by attractive Insecticide-treated Nets (ITNs) compared to inert ones and (B) by attractive and inert ITNs compared to deterrent ITNs for varied levels of physiological resistance in the vector population (feedbing attempt survival, 1 − µ_p1_). *Vector preference p*_*ITN*_ *of 0*.*6 indicates that 60% of malaria vectors choose an ITN-protected human if presented against an unprotected human. The reference inert ITN (panel A) induces a vector preference of 0*.*5 (vectors are equally likely to select an ITN-protected or unprotected human) and the reference deterrent ITN (panel B) induces a vector preference of 0*.*36 (only 36% of the vectors choose an ITN-protected human against an unprotected human). Lines become grey in the range [82; 100%], corresponding to survival levels that were never observed in* [12] *(supplementary Figure 2)*.

### Effect of quantitative behavioral resistance (escape behavior)

We then analyzed the impact of quantitative behavioral resistance on the efficacy of attractive ITNs. Specifically, we simulated increasing levels of indoor escape behavior by varying the diversion probability (*D*_*p*_) in our model (Figure 1). A diverted mosquito postpones host-seeking until the following night. The simulations demonstrated that, regardless of the level of indoor escape, attractive ITNs consistently outperform inert and deterrent ITNs in reducing malaria transmission. However, as indoor escaping increases, the relative efficacy of attractive ITNs decreases (Figure 4). When compared to an inert ITN (Figure 4A), a moderately attractive ITN is expected to reduce malaria transmission by 21% (under high resistance, i.e. 51% diversion that is the highest level of diversion observed in field data [12]; supplementary Figure 2B) to 30% (under no resistance, 0% diversion), while a highly attractive ITN is expected to reduce transmission by 39 (high resistance, 51% diversion) to 51% (no resistance, 0% diversion). When compared to a deterrent ITNs (Figure 4B), malaria transmission reduction ranged from 28 to 39% for inert ITN, from 43 to 57% for moderately attractive ITNs (*p*_*ITN*_ = 0.6) and from 56 to 70% for highly attractive ITNs (*p*_*ITN*_ = 0.7), depending on the level of resistance (i.e. between 0 and 51% diversion). Compared to no nets (0% use), attractive LLINs (50% use) are predicted to achieve a 88% reduction in transmission potential under high resistance conditions (51% diversion), which is 15 percentage points (p.p.) more than deterrent ITNs (supplementary Figure 3C).

**Figure 4.**
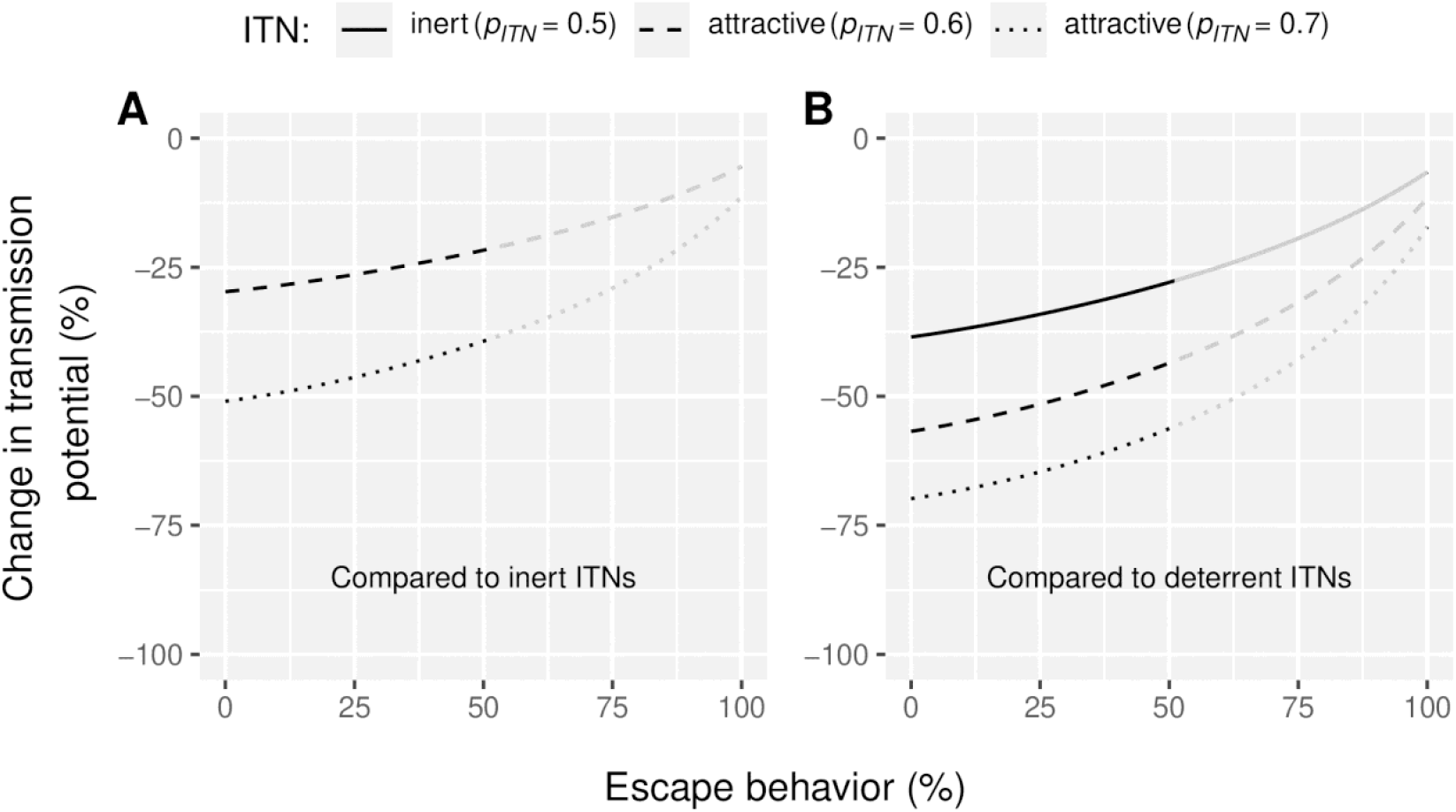
Reduction in *Plasmodium* transmission potential (*RedTP*) induced (A) by attractive Insecticide-treated Nets (ITNs) when compared to inert ones and (B) by attractive and inert ITNs when compared to deterrent ITNs for varied levels of quantitative behavioral resistance (escape behavior expressed as percent diversion, *D*_*p*_) in the vector population. *Vector preference p*_*ITN*_ *of 0*.*6 indicates that 60% of malaria vectors choose an ITN-protected human if presented against an unprotected human. The reference inert ITN (panel A) induces a vector preference of 0*.*5 (vectors are equally likely to select an ITN-protected or unprotected human) and the reference deterrent ITN (panel B) induces a vector preference of 0*.*36 (only 36% of the vectors choose an ITN-protected human against an unprotected human). Lines become grey in the range [51; 100%], corresponding to diversion levels that were never observed in* [12] *(supplementary Figure 2)*.

### Effect of qualitative behavioral resistance (spatiotemporal avoidance)

Mosquito vectors of malaria can alter their behavior to seek blood meals at different times or locations (i.e. outdoors), reducing their exposure to ITNs. We simulated this spatiotemporal avoidance of ITNs by varying the parameter Π, which represents the proportion of normal exposure to mosquito bites upon humans lacking ITNs, which occurs indoors at times when nets would normally be in use. In other words, Π quantifies the fraction of total bites that ITNs effectively prevent. As spatiotemporal avoidance increases, Π decreases. Regardless of the level of spatiotemporal avoidance, attractive ITNs consistently reduce malaria transmission compared to both inert and deterrent ITNs (Figure 5). However, as spatial-temporal avoidance increases, the relative efficacy of attractive ITNs decreases. At 62% spatial-temporal avoidance (i.e. the highest value reported in literature [31]), attractive ITNs are expected to reduce malaria transmission by 17 and 31% compared to inert ITNs (Figure 5A), for moderately and highly attractive ITNs, respectively, and by 35 and 46% compared to deterrent ITNs (Figure 5B). Under the same conditions (62% spatiotemporal avoidance), an inert ITN is expected to reduce malaria transmission by 22% when compared to a deterrent ITN (Figure 5B). Compared to no nets (0% use), attractive LLINs (50% use) are predicted to achieve a 90% reduction in transmission potential under high resistance conditions (62% spatiotemporal avoidance), which is 14 percentage points (p.p.) more than deterrent ITNs (supplementary Figure 3D).

**Figure 5.**
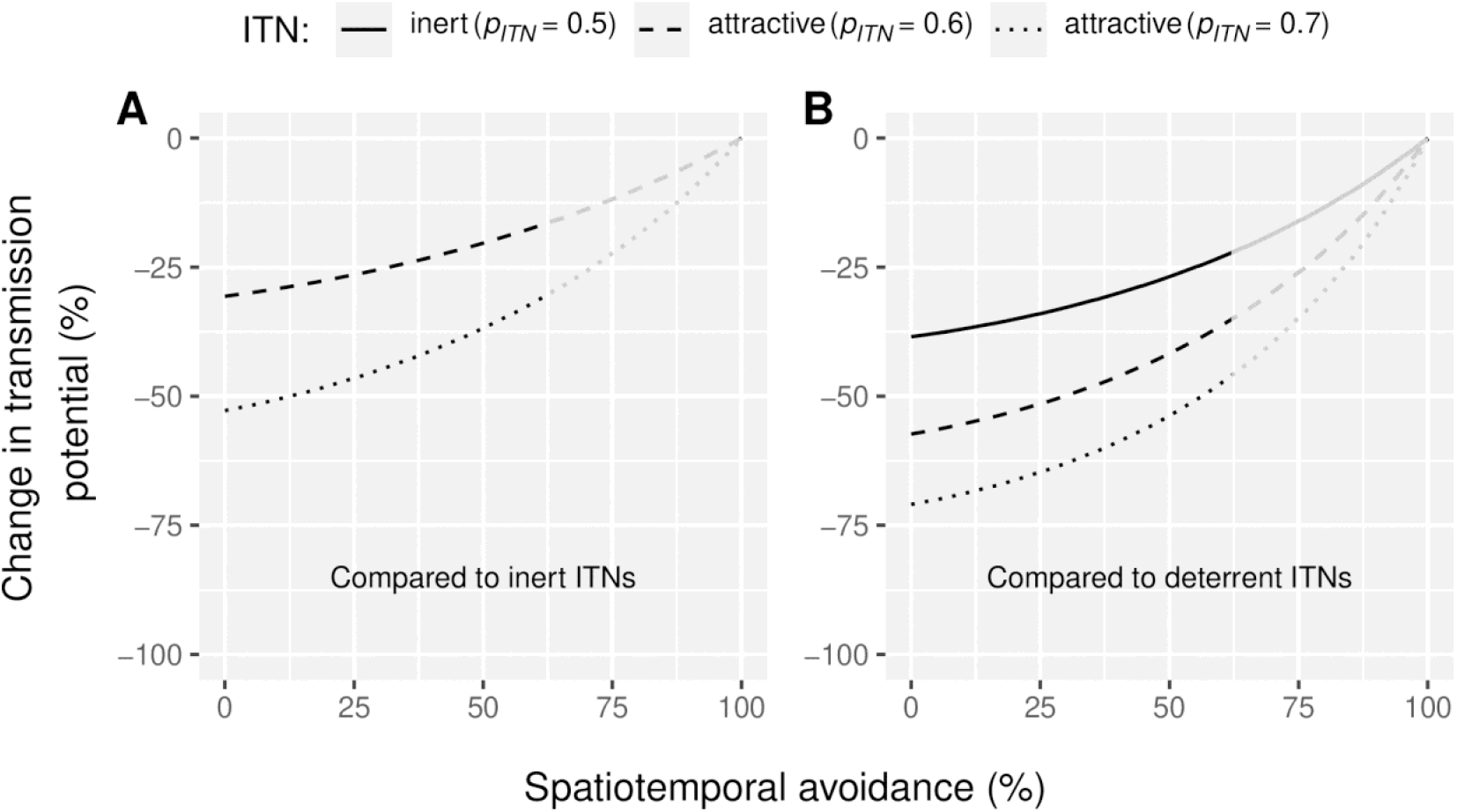
Reduction in *Plasmodium* transmission potential (*RedTP*) induced (A) by attractive Insecticide-treated Nets (ITNs) when compared to inert ones and (B) by attractive and inert ITNs when compared to deterrent ITNs for varied levels of qualitative behavioral resistance (spatiotemporal avoidance expressed as 1 minus the efficacy of ITNs in reducing the number of bites, 1 − Π) in the vector population. *Vector preference p*_*ITN*_ *of 0*.*6 indicates that 60% of malaria vectors choose an ITN-protected human if presented against an unprotected human. The reference inert ITN (panel A) induces a vector preference of 0*.*5 (vectors are equally likely to select an ITN-protected or unprotected human) and the reference deterrent ITN (panel B) induces a vector preference of 0*.*36 (only 36% of the vectors choose an ITN-protected human against an unprotected human). Lines become grey in the range [62; 100%], corresponding to spatiotemporal avoidance levels that were never observed in the field (see refs for parameter* Π *in Table 1)*.

### Uncertainty and sensitivity analyses

The uncertainty analysis revealed that, when all parameters varied within their plausible ranges, the mean reduction in transmission potential with attractive ITNs was estimated at 39.3% (95% Credible Interval: [11.9%, 71.4%]) relative to deterrent ITNs. The sensitivity analysis (Figure 6) identified the parameters with the greatest influence on model outcomes among those not expressly analyzed (Figure 6). Notably, we found that the reduction in transmission potential was greater for *Anopheles* populations with a higher baseline daily survival probability (*S*; Partial Rank Correlation Coefficients PRCC= −0.54 [95% CI: −0.63, −0.48], Partial Slope Coefficient PSC= −40.0 [95% CI: −44.7, −35.6]), shorter gonotrophic cycle duration (*g* in days; PRCC= 0.67 [0.32, 0.72], PSC= 5.76 [5.41, 6.19]), higher post-bite mortality probability in rooms with ITNs (*µ*_*2p*_; PRCC= −0.55 [−0.63, −0.48], PSC= −15.6 [−17.7, −13.2]), and when parasites exhibited longer extrinsic incubation period duration (*n* in days; PRCC = −0.48 [−0.55, −0.41], PSC = −1.68 [: −1.91, −1.41]).

**Fig 6:**
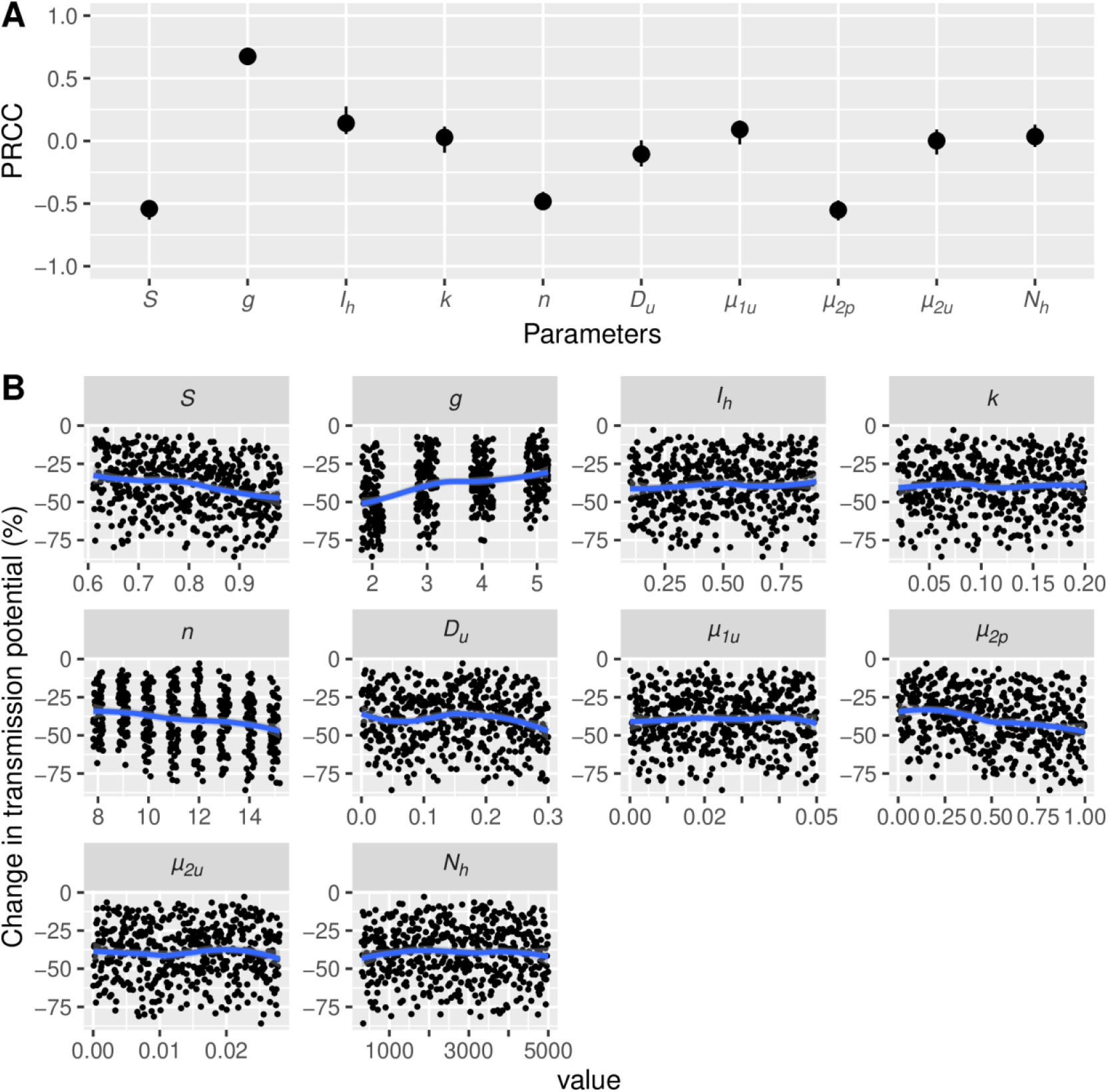
Results of a sensitivity analysis using Latin Hypercube Sampling, showing (A) Partial Rank Coefficients of Correlation (PRCC) of parameters and (B) simulation plots illustrating the impact on transmission potential of varying parameter values when comparing a moderately attractive ITN (*p*_*ITN*_ = 0.6) to a deterrent ITN (*p*_*ITN*_ = 0.36). *Intervals in panel A are 95% Confidence Interval of PRCC. In panel B, percent change in transmission* potential (*RedTP*) *is plotted against values for each model parameter (x-axis). Blue curves on panel B are loess regression. See Table 1 for parameters’ definitions*.

## Discussion

Recent evidence from both field and laboratory studies suggests that some ITNs may increase the attraction of *An. gambiae* mosquitoes to human hosts, potentially raising the bite exposure of ITN users [12,32]. However, the implications of such attractive ITNs on malaria transmission at the community level remained unclear. To address this, we developed a model to assess their impact under various levels of ITN usage and vector resistance, including both physiological and behavioral resistance.

Our model predicts that attractive ITNs consistently reduce malaria transmission across all simulated scenarios compared to inert or deterrent ITNs, even when lower ITN usage rates and the presence of physiological or behavioral resistance limits the magnitude of the effect. Sensitivity analysis identified four additional key parameters that significantly influence the relative efficacy of attractive ITNs: daily survival probability (*S*), gonotrophic cycle duration (*g*), the extrinsic incubation period (*n*), and post-bite mortality in the room of an ITN-protected individual (*µ*_*2p*_). The general pattern is that any factor reducing the average infectious life expectancy of mosquitoes, whether through increased mortality or more frequent encounters with ITNs, tends to enhance the relative efficacy of attractive ITNs. The corollary is that shorter extrinsic incubation periods reduce their relative efficacy, since parasites can reach transmissibility within a shorter mosquito lifespan.

In all cases, attractive ITNs performed better than inert ITNs, which in turn outperformed deterrent ITNs in reducing malaria transmission. Currently, most widely distributed ITNs have deterrent properties [12,26]. Our findings suggest that shifting from deterrent to inert (or attractive) ITNs could substantially enhance malaria control efforts, as also suggested by Killeen *et al*. [11]. We therefore urge national and international organizations responsible for ITN evaluation, selection, and distribution, in collaboration with manufacturers, to reassess whether deterrence should remain a desirable characteristic of ITNs.

Increasing reports of attractive effects in ITNs [12] should not be seen as a drawback but rather as an opportunity for innovation in malaria control. Our findings suggest that attractive ITNs could significantly enhance malaria prevention, opening new avenues for product development. Recent advances in mosquito chemical ecology have identified numerous host-derived attractants [33], while ITN manufacturers have developed techniques to incorporate dual compounds, such as insecticides and synergists [34]. Leveraging these innovations, next-generation ITNs could be designed to integrate both insecticidal and attractive properties, thereby optimizing vector control strategies. Such innovations could enhance the effectiveness of ITNs while maintaining their widespread acceptance and use in malaria-endemic regions. One key advantage of combining insecticides and attractants into ITNs is their potential to remain effective even in the absence of humans in the room or when old ITNs are repurposed for alternative uses [35]. This approach could extend the operational lifespan and functional utility of ITNs, further strengthening their role in malaria control. Nevertheless, it is important to recognize the potential adverse effects of attractive ITNs. By increasing host attractiveness, such nets could expose users to higher biting risk in situations of incomplete coverage or physical deterioration, which may reduce user satisfaction and compliance. These challenges underscore that, alongside technical optimization, the success of attractive ITNs would also depend on strong community engagement and communication strategies that emphasize their communal benefits while mitigating concerns about personal protection and nuisance biting [11,12].

While our model provides valuable insights into the potential impact of attractive ITNs on malaria transmission, it is important to acknowledge its limitations. The model relies on simplified assumptions about mosquitoes, parasites, and humans, which may limit its applicability to all real-world settings. In particular, we assume binary host choice, with mosquitoes selecting between ITN-protected and unprotected humans, without accounting for alternative hosts such as livestock. In areas where alternative hosts are abundant, mosquito feeding patterns could shift [36,37]. Our model also assumes spatio-temporal homogeneity, meaning that human and mosquito populations are evenly mixed in space and that model parameters remain constant over time (i.e., the model is static). This simplification neglects potential spatial heterogeneity, such as clustering of hosts, as well as temporal variability driven by seasonality, intervention effects, or evolutionary processes. Important model parameters such as daily survival probability, gonotrophic cycle duration, and the extrinsic incubation period are strongly driven by environmental factors, either directly (e.g., temperature and humidity [28,38,39]) or indirectly through their dependence on favorable ecological conditions determining the spatio-temporal distribution of the mosquito. It is therefore reasonable to expect the relative efficacy of attractive ITNs to vary across ecological settings.In variable real-world settings, such heterogeneities can influence both mosquito–human interactions and malaria transmission dynamics [8,40–42]. Another assumption is linked to the deterministic nature of our model, which does not account for stochastic variability in input parameters. Stochastic effects may be particularly relevant in small populations or low-transmission settings, where random events can substantially influence outcomes. Relaxing these assumptions could alter quantitative predictions, potentially amplifying or dampening the effects of attractive ITNs in specific settings. However, our uncertainty analysis, varying all parameters across plausible ranges, indicates that attractive ITNs consistently reduce transmission potential more than deterrent ITNs under all simulations. This suggests that the relative benefit of attractive ITNs is likely robust, even if the absolute magnitude of their impact could vary in more complex scenarios. Incorporating these complexities into future modeling efforts could refine predictions and improve the relevance of our findings for diverse transmission settings.

As with any model that has not been validated using field data, these results should be interpreted with caution and considered to have theoretical value only. Validation of our model predictions would ideally require cluster-randomized controlled trials using ITNs whose attractiveness, inertness, or deterrence has been assessed beforehand in the vector population to ensure the interventions accurately reflect their intended behavioral properties.

## Conclusion

Our findings predict that attractive ITNs could significantly reduce malaria transmission at the community level, even in the presence of physiological or behavioral resistance in vectors, relative to inert and deterrent ITNs. This challenges the conventional preference for deterrent ITNs, which prioritize personal protection, and expands the possibilities for designing next-generation ITNs. By combining insecticidal and attractant properties, innovative ITNs could enhance malaria control by remaining effective despite high resistance levels and repurposing. Advances in mosquito chemical ecology and ITN manufacturing present a unique opportunity to develop more efficient tools for malaria prevention. Future research should focus on better understanding the mechanisms underlying the attractant effect of some current ITNs, validating the theoretical predictions through field trials, and fostering collaborations with manufacturers to translate these insights into practical ITN prototypes.

## Data Availability

All data produced are available online at https://doi.org/10.5281/zenodo.15121768

## Funding

The authors declare that they have received no specific funding for this study.

## Conflict of interest disclosure

The authors declare that they comply with the PCI rule of having no financial conflicts of interest in relation to the content of the article. The authors are recommenders for PCI Zoology. CP is recommender for PCI Infection.

## Author contributions

Conceptualization: NM, CP; Methodology: NM; Software: NM; Formal analysis: NM; Investigation: NM, CP; Data curation: NM; Writing – original draft: NM; Writing – review and editing: NM, CP. Visualization: NM.

## Data, scripts and code availability

Scripts, code and data are available online: https://doi.org/10.5281/zenodo.15121768 [43].

## Supplementary information

Supplementary information is available online: https://doi.org/10.5281/zenodo.15121768 [43].

## Notes

### Competing Interest Statement

The authors have declared no competing interest.

### Funding Statement

This study did not receive any funding

### Summary of Updates

This is the peer-reviewed and recommended version of the preprint (https://doi.org/10.24072/pci.infections.100244). PCI Infections badge added. Line numbering removed.

