## Supplementary material for "The potential of attractive insecticide-treated nets (ITNs) in reducing malaria transmission: a modeling study": Supplmentary Figures 1, 2, & 3

### Supplementary materials

Supplement to: Moiroux N, Pennetier C. The potential of attractive insecticide-treated nets (ITNs) in reducing malaria transmission: a modeling study [Internet]. medRxiv; 2025. https://doi.org/10.1101/2025.04.02.25325102.

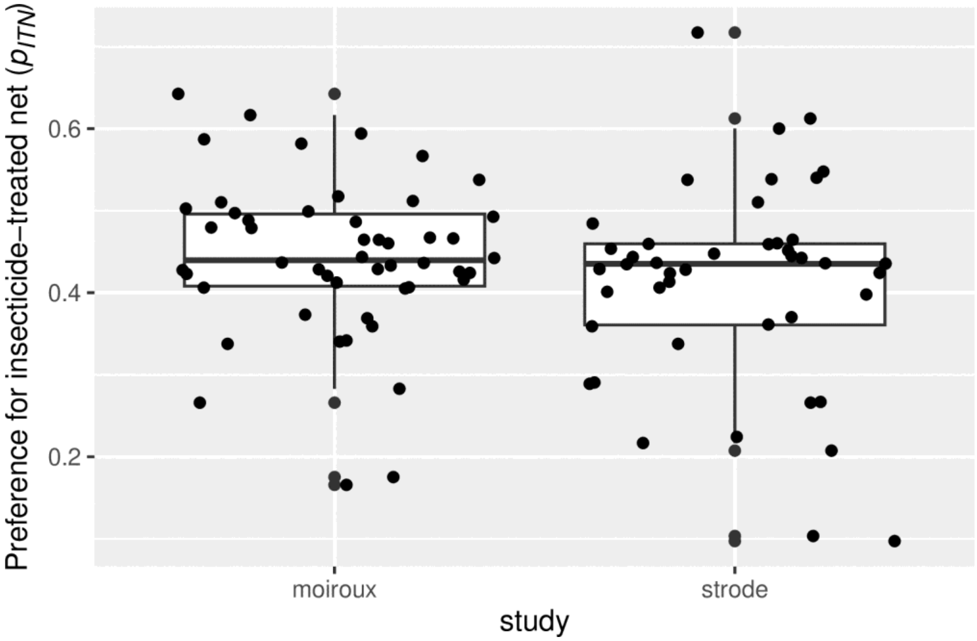


#### Supp. Figure 1: Vector preference for insecticide treated nets measured in the fields through experimental hut trials.

*Data come from two meta-analysis studies* (Strode *et al.*, 2014; Moiroux *et al.*, 2017). *Each dot represent an ITN. Boxplots show 25$^{th}$, 50$^{th}$, and 75$^{th}$ percentiles, whiskers indicate 5$^{th}$ and 95$^{th}$ percentiles. There is 50 values coming from 9 Experimental hut Trials (EHT) in Moiroux et al. and 44 points coming from 13 EHT in Strode et al. Most of points are below 0.5 indicating deterrence. About 25 % of points are between 0.5 and 0.75 indicating attraction.*


Moiroux, N. *et al.* (2017) 'Remote Effect of Insecticide-Treated Nets and the Personal Protection against Malaria Mosquito Bites', *PLOS ONE*, 12(1), p. e0170732. Available at: https://doi.org/10.1371/journal.pone.0170732.

Strode, C. *et al.* (2014) 'The Impact of Pyrethroid Resistance on the Efficacy of Insecticide-Treated Bed Nets against African Anopheline Mosquitoes: Systematic Review and Meta-Analysis', *PLoS Med*, 11(3), p. e1001619. Available at: https://doi.org/10.1371/journal.pmed.1001619.

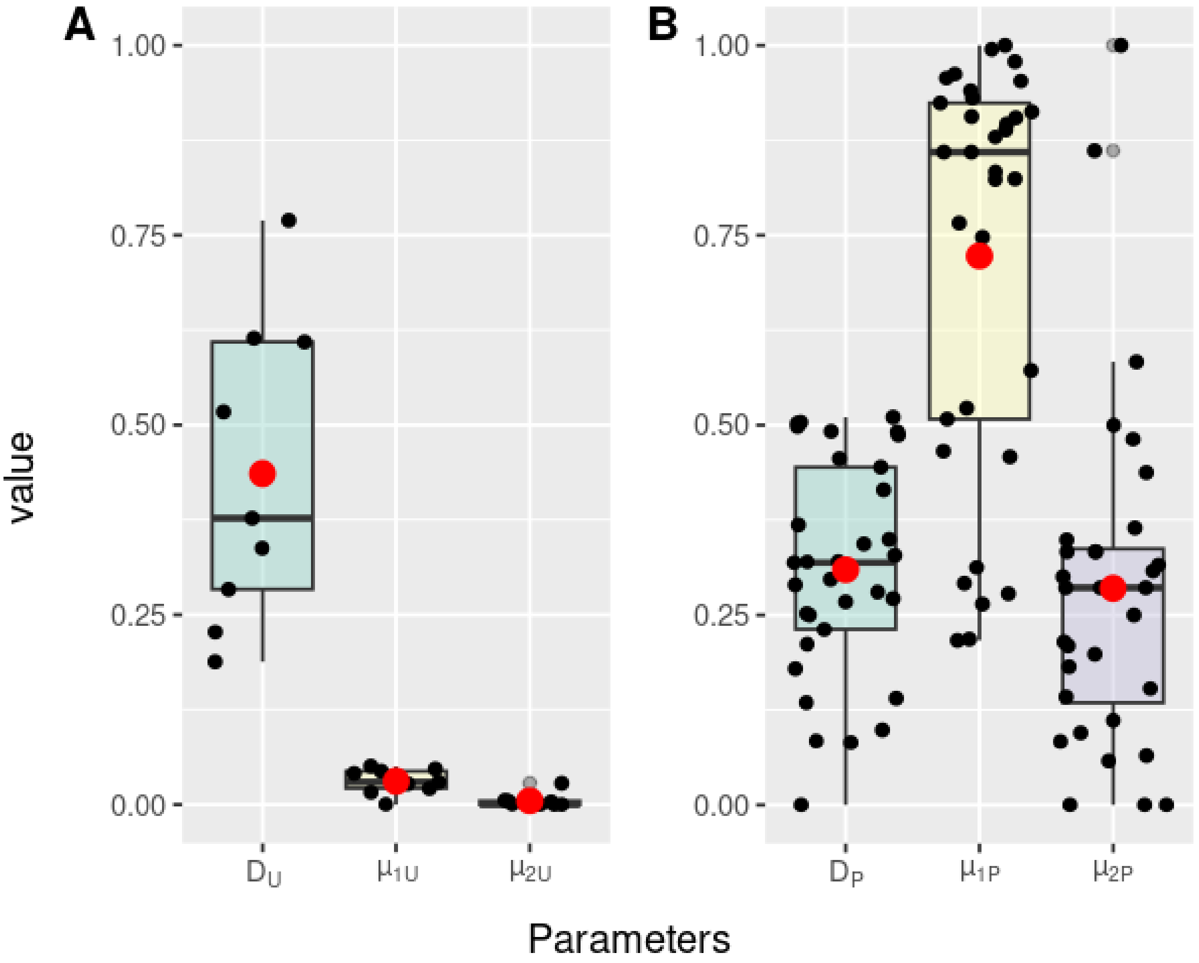


Supp. Figure 2: Fields measure of diversion ($D$), pre-bite mortality ($\mu_1$) and post-bite mortality ($\mu_2$) in an experimental hut with (A) an unprotected human or (B) an ITN-protected human.

*Red dots indicates arithmetic means that were used as default value in the model.*

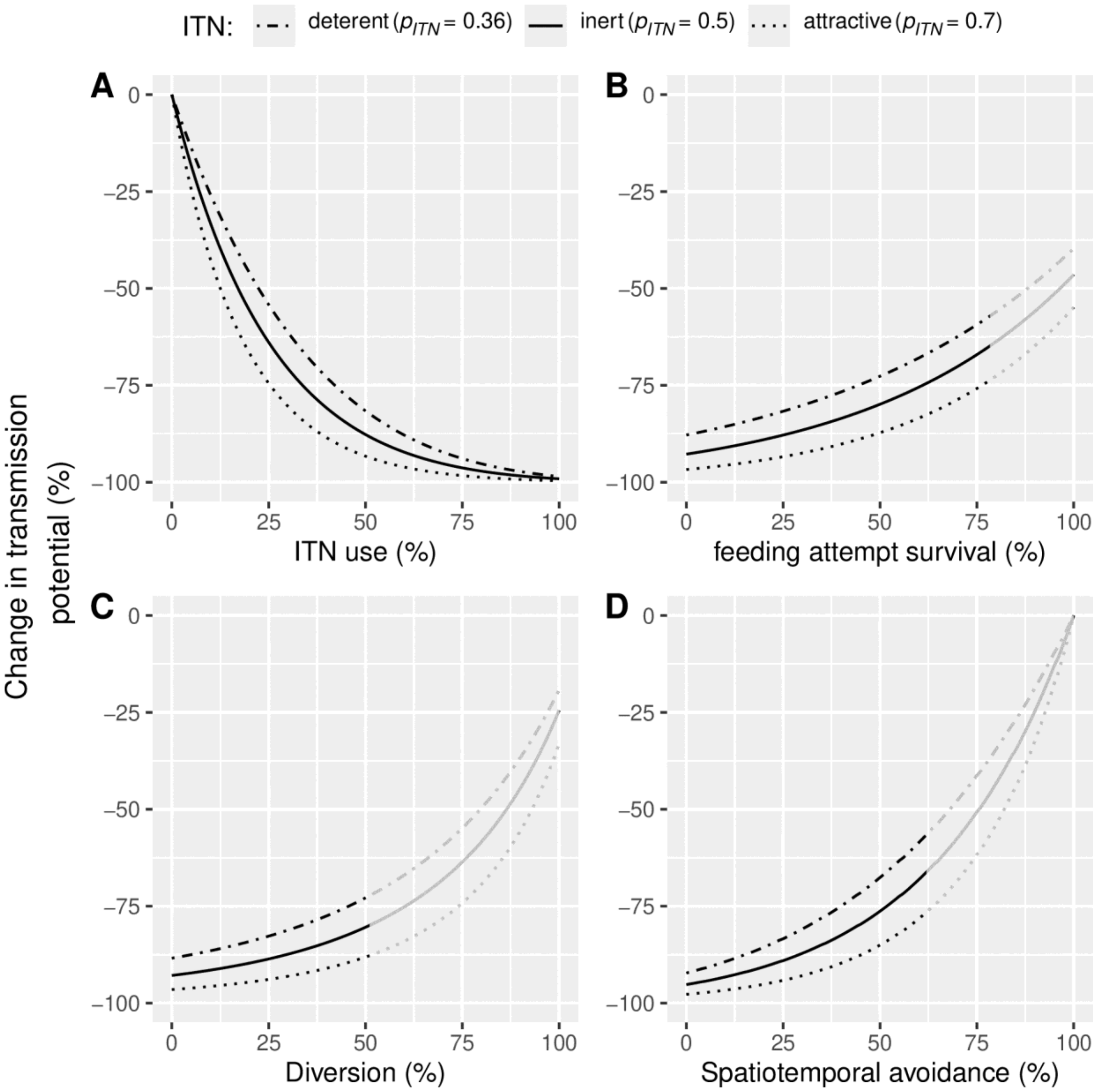


Supp. Fig 3: Reduction in *Plasmodium* transmission potential induced by deterrent, inert, and attractive insecticide-treated nets (ITNs), compared to no nets, for varied levels of (A) ITN usage rates, (B) physiological resistance, (C) quantitative behavioral resistance, and (D) qualitative behavioral resistance.

*Lines become grey in ranges of values that were never observed in [12] (supplementary Figure 2).*
